# Efficacy of Improvised Cervical Immobilization Techniques in Remote Settings: A Systematic Review

**DOI:** 10.1101/2025.07.14.25331399

**Authors:** Inam S. Ahmed

**Affiliations:** Independent Researcher and Wilderness Medicine Practitioner

## Abstract

**Introduction:** Cervical spine injuries in wilderness and austere environments present unique challenges due to the lack of commercial immobilization devices and prolonged evacuation times. Improvised cervical collars—made from items such as SAM® splints, rolled towels, or fleece garments—are frequently used in these settings, yet their biomechanical effectiveness and field practicality have not been systematically reviewed. This article synthesizes current evidence on the efficacy, safety, and feasibility of improvised cervical immobilization techniques used in remote environments.

**Method:** We conducted a systematic review in accordance with PRISMA guidelines, searching five databases (PubMed, Scopus, Web of Science, CINAHL, and Google Scholar) for studies published between January 2000 and March 2025. Inclusion criteria encompassed biomechanical or qualitative evaluations of improvised cervical immobilization methods in human volunteers or cadaveric models, with relevance to wilderness, prehospital, or austere settings. Eleven studies met inclusion criteria.

**Results:** Findings suggest that improvised collars particularly SAM splints and rolled fleece or towel collars can reduce cervical motion to levels comparable to standard rigid collars in most planes of movement. Spray-foam devices demonstrated even greater stability in cadaveric models. Soft improvised collars consistently outperformed foam commercial collars in both stability and comfort. Improvised methods also showed favorable usability in the field, though their application requires proper training.

**Conclusion:** Improvised cervical spine immobilization techniques can be biomechanically effective and operationally valuable in remote care contexts. When rigid collars are unavailable, well-applied improvised devices offer a viable alternative and should be included in wilderness medicine education. Further field-based research is warranted to evaluate real-world outcomes.

## INTRODUCTION

Spinal injuries in wilderness or austere settings pose unique challenges. Standard prehospital practice has long involved rigid cervical collars and backboards, but evidence supporting these measures is limited ^5^. In fact, growing data suggest that traditional immobilization may cause harm (e.g., airway compromise, increased intracranial pressure, pressure ulcers) without proven benefit.^6, 8^ Modern guidelines emphasize spinal motion restriction (SMR) over rigid immobilization, focusing on gentle handling and patient cooperation rather than forceful immobilization.^6, 8^ The Wilderness Medical Society’s 2023 guidelines explicitly state there is “no requisite role for commercially made or improvised rigid cervical collars in any out-of-hospital environment,” favoring soft collars or padding if needed.^8^ Similarly, many EMS systems now use selective SMR protocols, often replacing hard collars with soft collars or none in certain cases.^6^

In wilderness contexts, rescuers rarely carry bulky commercial cervical collars or full backboard kits. Instead, they must improvise with available materials (e.g. clothing, camping gear). Common makeshift cervical spine supports include rolled towels or blankets, rolled jackets/ fleece garments, SAM® splints molded as collars, tape-and-padding arrangements, or newer concepts like spray-foam splints. These improvised collars aim to reduce neck motion until evacuation, but their effectiveness and safety require scrutiny. Questions remain about how well improvised devices limit cervical motion compared to standard devices, and what trade-offs exist in comfort, practicality, and risk. Additionally, rescue personnel must be trained to apply improvised collars correctly under field conditions, all while balancing patient safety (e.g. avoiding airway obstruction or carotid compression) and the real risk of prolonged evacuation times.

This review synthesizes available evidence on improvised cervical spine immobilization in wilderness, austere, or remote settings. We expand on our work by identifying studies including biomechanical trials (human volunteer and cadaveric), field evaluations, and relevant guidelines from wilderness, military, and EMS domains. We focus on five key considerations: biomechanical efficacy (motion restriction achieved), field practicality (availability and ease of use), training requirements for proper application, patient safety (risks/benefits), and device limitations under real-world conditions. By consolidating current knowledge on makeshift cervical collars from rolled jackets and SAM splints to foam-based splints, we aim to inform best practices for wilderness spinal injury management and highlight gaps for future research.

## METHODS

### Search Strategy

We conducted a systematic literature search in five electronic databases: PubMed, Scopus, Web of Science, CINAHL, and Google Scholar. The search spanned from January 1, 2000 through March 31, 2025, capturing studies published in this period. We used combinations of keywords related to cervical spine immobilization and austere or wilderness settings. Search terms included “cervical spine”, “improvised collar”, “SAM splint”, “towel roll”, “wilderness medicine”, “austere immobilization”, “spray-on foam”, and “prehospital spinal care”. These terms were entered in various Boolean combinations to ensure broad coverage. No language restrictions were applied during the search except that only English-language results were considered at the screening stage. In addition to the database queries, we examined the reference lists of relevant articles and utilized Google Scholar to identify grey literature or any additional reports that might have been missed by the indexed database search.

### Eligibility Criteria

Studies identified through the search were screened against predefined inclusion and exclusion criteria. Inclusion criteria were defined a priori as studies that met all of the following:

- Evaluated improvised cervical immobilization techniques (e.g., makeshift collars or splints) in human adult volunteers or cadaveric models (simulated patients).
- Reported biomechanical outcomes (such as cervical range of motion restriction or stabilization) or qualitative assessments (such as comfort, feasibility, or practical effectiveness) of the immobilization technique.
- Conducted in wilderness, austere, prehospital EMS, or other remote settings, or explicitly addressing challenges of such environments.
- Published in English in peer-reviewed literature.

Exclusion criteria were applied to filter out:

- Studies set exclusively in urban or hospital environments without relevance to remote or resource-limited settings.
- Trials examining only commercial cervical collars or devices (with no improvised component evaluated).
- Studies focusing solely on pediatric populations (children/adolescents), since pediatric cervical spine management presents different considerations and was beyond this review’s scope.
- Articles not in English, or those for which full-text was unavailable or did not contain original data (e.g., editorials, commentaries).

### Study Selection

All references identified through the searches were imported into a reference management software, and duplicate records were removed. The remaining unique citations then underwent a two-stage screening process. In the first stage, two reviewers independently screened titles and abstracts to determine potential relevance based on the eligibility criteria. Studies that clearly did not meet inclusion criteria (for example, wrong population or intervention, or not related to cervical immobilization) were excluded at this stage. In the second stage, we obtained the full-text articles for all remaining references deemed potentially eligible. Each full text was assessed in detail by the same two reviewers against the inclusion/exclusion criteria. Any disagreements or uncertainties in study selection were resolved through discussion and consensus, with a third reviewer available to adjudicate if needed.

We recorded the study identification and selection process in a PRISMA flow diagram (Figure 1). Figure 1 illustrates the number of records retrieved, screened, and ultimately included. In total, our search yielded approximately 220 records across all sources. After removing duplicates, 168 unique records remained. Of these, 168 were screened by title/abstract, and ∼140 were excluded as clearly irrelevant or not meeting criteria. We assessed 28 full-text articles for eligibility. Following full-text review, 17 studies were excluded for reasons such as improper setting (e.g., purely hospital-based studies), intervention not improvised (commercial collar only), or outcomes not pertinent to immobilization efficacy. Ultimately, 11 studies met all inclusion criteria and were included in the qualitative synthesis. The included studies comprised a mix of experimental designs, including volunteer trials in simulated wilderness settings and cadaveric biomechanical studies, reflecting the limited but emerging evidence base on improvised cervical spine immobilization.

**Figure 1.**
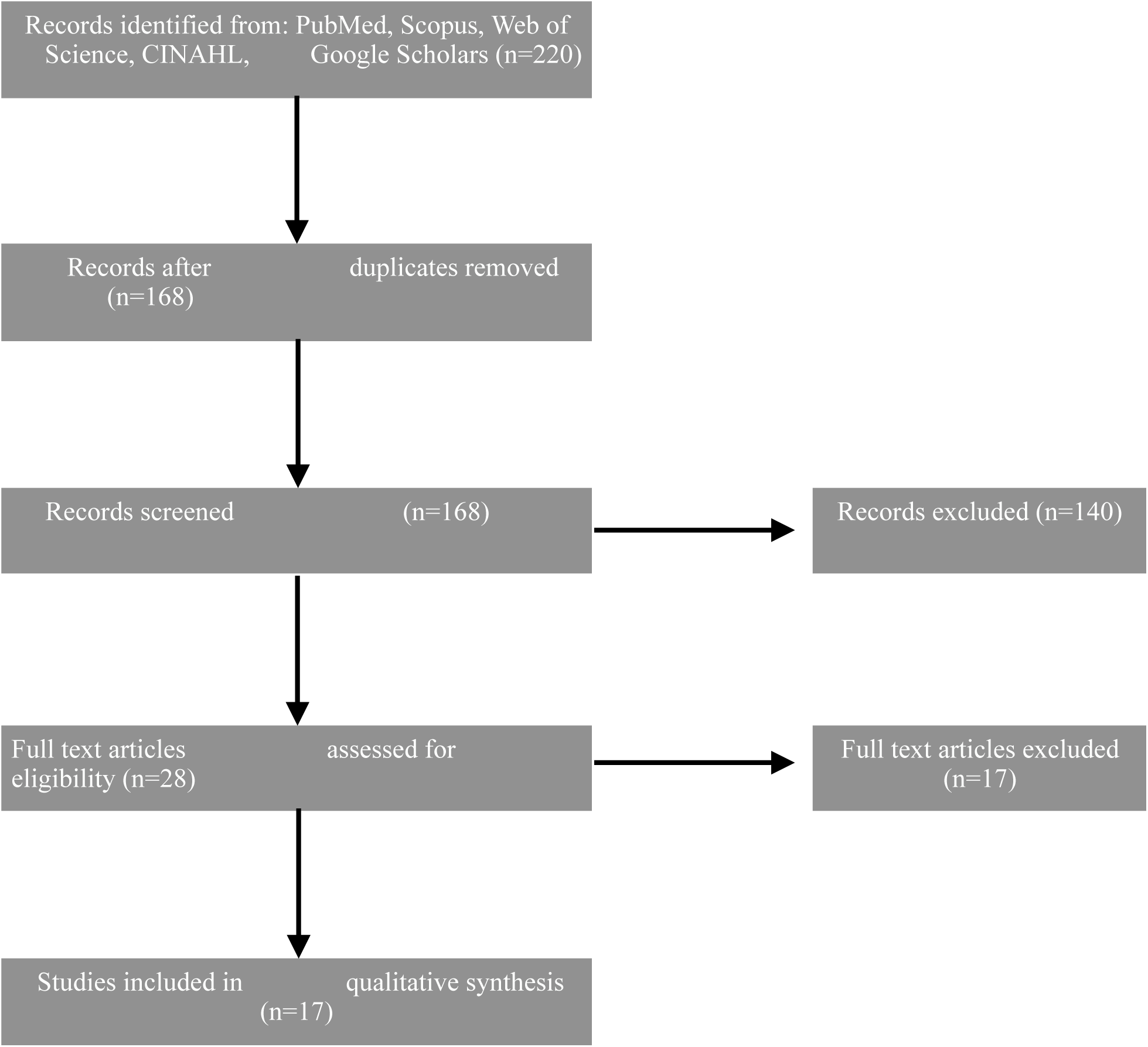
PRISMA flow diagram: 220 records identified; 168 screened; 11 studies included.

### Data Extraction and Quality Assessment

For each of the 11 included studies, two reviewers independently extracted data using a standardized form. The extracted data elements included: study design and setting (wilderness/ austere field simulation, laboratory, etc.), sample characteristics (number of subjects and whether healthy volunteers or cadavers were used), details of the improvised immobilization technique evaluated (e.g., SAM® splint molded as a collar, rolled blankets or towels, spray-foam splinting products, etc.), the comparator or control condition if applicable (such as standard rigid cervical collar or no immobilization), and the outcome measures reported. Key outcomes were typically biomechanical (degree of cervical motion restriction in various planes, stability under movement or load, etc.), though some studies also reported qualitative outcomes like user comfort or application time. Any clarification needed from study authors was planned via contact if data were unclear, though in practice all necessary information was obtained from the publications.

We also appraised the methodological quality and risk of bias of the included studies. Given the heterogeneity of study types, we did not use a single instrument across all studies; instead, we tailored our quality assessment to each study design. For example, for controlled laboratory trials with volunteer participants, we examined elements akin to risk-of-bias criteria (randomization or repeated-measures design, blinding of outcome measurement, etc.). For cadaveric or device simulation studies, we assessed factors such as robustness of the experimental setup, sample size, and potential conflicts of interest. We noted common limitations, including small sample sizes and lack of blinding (inherent in many device comparison studies), and we qualitatively judged whether these might introduce bias. Two reviewers performed the quality assessments independently, and any differences in judgments were resolved by consensus. No study was excluded on the basis of quality; however, the strength of evidence was considered when drawing conclusions, and study limitations are acknowledged in our analysis.

### Data Synthesis

We planned a descriptive, narrative synthesis of the findings because the included studies were diverse in methods and outcome metrics, precluding a meaningful quantitative meta-analysis. Results from each study were organized and summarized to facilitate comparison across different improvised immobilization techniques. We grouped the evidence by the type of improvised method: for example, soft improvised collars (such as those made from clothing or towels), structural splints (like SAM splints molded to the neck), and novel materials (such as one-step spray-on foam splints). For each category, we qualitatively compared the degree of cervical spine motion restriction achieved relative to standard care (commercial rigid collars or no immobilization) and highlighted any notable advantages or challenges reported. This synthesis approach allowed us to integrate biomechanical outcomes (e.g. measured reduction in range of motion) with practical considerations (e.g. comfort, ease of application in the field) across studies. All data synthesis and interpretations were done by at least two authors working together to ensure accuracy and consistency.

## RESULTS

### Biomechanical Efficacy of Improvised vs Standard Collars

Rolled Jacket (Fleece) vs Rigid Collar: Porter et al. (2019) conducted a volunteer trial in a backcountry simulation comparing a fleece jacket improvised collar to a standard rigid C-collar.^10^ Twenty-four healthy subjects were tested for neck range of motion in flexion/extension, lateral bending, and rotation with each collar type, using a goniometer. The improvised fleece collar was found non-inferior to the commercial rigid collar in limiting cervical motion across all planes. In other words, the rolled fleece jacket provided essentially comparable restriction of movement as a traditional hard collar. Notably, participants rated the comfort significantly higher with the fleece collar. The authors concluded that “an improvised fleece jacket collar” can be an effective field substitute for a standard C-collar in suspected spine injury during wilderness care.

SAM Splint vs Philadelphia Collar: An earlier study by McGrath and Murphy (2009) specifically evaluated a SAM® splint molded into a cervical collar shape versus a conventional hard Philadelphia collar.^11^ In a brief report with healthy volunteers, cervical range of motion was measured with each device. The SAM splint collar was found to be as effective as the Philadelphia collar at restricting neck movement. The authors reported no significant difference in the degree of motion limitation between the malleable aluminum-foam splint (once properly molded and secured around the neck) and the rigid commercial collar. This suggests a SAM splint. a common item in wilderness first aid kits can function as a credible cervical immobilizer when shaped and applied correctly.

Fig. 2: Molding a SAM splint into an improvised cervical collar. A study found a SAM splint collar as effective as a standard Philadelphia hard collar in limiting cervical spine motion.^11^

Folded Towels vs Rigid Collar: Beyond fleece and SAM splints, a recent non-inferiority trial by Eisner et al. (2022) examined folded towels as a low-cost immobilization method in comparison to standard rigid collars and foam cervical collars.^1^ Thirty healthy volunteers were tested using an electronic cervical range-of-motion device in six directions, both seated and supine. A folded bath towel was wrapped circumferentially around the neck and crossed over the chest (secured under the arms) as the improvised support.^14^ The rigid collar reduced median cervical motion by about 36.8° (seated) relative to no immobilization, whereas the folded towel reduced motion by ∼27.0° (seated). Using a composite motion score, folded towels met the pre-defined non-inferiority margin compared to the hard collar (composite scores 0.89 seated and 0.47 supine, where <1.0 indicates non-inferior). In practical terms, towels achieved roughly 75% of the motion restriction of a rigid collar, being particularly effective at limiting extension and rotation. However, the towel was less effective for flexion (forward bending), which remained a relative weak point. By contrast, a soft foam cervical brace (commercial soft collar) performed significantly worse – it reduced motion by only ∼14° and failed non-inferiority, indicating inferior stabilization. The authors concluded that folded towels are a viable non-inferior option for cervical spine immobilization in resource-limited settings, especially when combined with proper patient positioning (e.g., supine on a firm surface). They suggested that towels could be paired with a backboard or litter to improve overall stability for transport.

Spray-On Foam vs SAM Splint (Cadaveric model): One innovative approach has been a spray-on foam splint (FastCast®) that hardens into a mold, proposed for military or austere use.

Roebke et al. (2023) performed a cadaveric study comparing this one-step spray foam collar to a SAM splint collar in truly unstable cervical injuries.^2^ Three cadaver specimens had C3–C4 destabilized (cervical corpectomy) to simulate a significant spine injury. Each cadaver’s neck was then immobilized with either a standard SAM splint collar or the FastCast spray foam (applied circumferentially around neck and head) in random order. Fluoroscopic imaging and orthopedic surgeon assessments were used to evaluate alignment maintenance before and after a log-roll and gravitational stress test. The results strongly favored the spray-on foam technique: 100% of FastCast applications maintained alignment after log-roll and under gravity stress, compared to only 33–47% of cases with the SAM splint (most SAM splints lost reduction or shifted). Orthopedic raters gave the FastCast significantly better immobilization scores than the SAM in all cases (median score ∼44 vs 32 out of 50; P < .01). The FastCast was thus rated superior to the SAM splint for stabilizing an unstable neck. Investigators noted the foam collar formed a custom fit, eliminated pressure points and did not require tight circumferential straps (improving airway and vascular access).

Other Devices: No published studies were found evaluating improvised use of duct tape alone or other ad-hoc materials in isolation. However, standard EMT practice has long used tape in conjunction with foam blocks or sandbags to secure the head to a backboard. In the absence of a backboard, wilderness protocols often involve using broad tape or straps to stabilize the head against a pad or stretcher. For instance, pediatric hospital guides note that rolled towels or IV fluid bags on either side of the head plus tape can substitute for head blocks when a collar alone is insufficient.^3,4^ The long-standing practice of sandbags and tape alone (without a collar) is generally not recommended due to poor restriction and potential for shifting.^5,6^ Thus, while duct tape is not a “cervical collar” per se, it is often a component of improvised immobilization e.g., to secure a rolled jacket in place or to remind the patient to keep still. No adverse events from using tape in this manner are reported in the literature, so long as care is taken never to encircle the neck and to monitor airway patency.

### Field Experience and Case Reports

Literature on actual field deployments of improvised c-spine immobilization is mostly limited to case discussions and training scenarios. Wilderness medicine courses (e.g., NOLS Wilderness Medicine) routinely teach improvised SMR. In a NOLS case study, a patient with suspected spine injury was packaged by first responders using a rolled-up fleece jacket as a cervical collar and clothing-stuffed head blocks, all secured to a foam sleeping pad.^6^ Upon handoff to an ambulance crew, the improvised collar was swapped for a commercial soft collar, and the patient was transported without a hard board or rigid collar, a practice that surprised the rescuers but reflects modern protocol. The case commentary highlighted that “paranoia and rigid immobilization are making way for careful handling and comfortable patient packaging” in current spine care, with language shifting from immobilization to protection. The authors noted that soft collars (including improvised ones) “prevent some movement and remind the patient and caregiver to be careful with the neck”. In fact, some EMS systems now favor soft collars over hard in many cases. The improvised fleece collar in this scenario was deemed to have “worked well” for the ∼20 minutes of waiting and was appropriate for a longer wilderness context if evacuation were delayed.

Another example in Wilderness & Environmental Medicine (WEM) described an alpine rescue where a jacket and stuff sacks were used to immobilize a climber’s head and neck until a litter evacuation was completed.^7^ These anecdotal reports underscore that improvised collars are being used successfully in the field, at least as an interim measure until definitive care or more formal equipment is available. However, systematic data on outcomes (e.g., neurologic deterioration or patient comfort during long evacuations) are lacking. Guidelines and Expert Recommendations

Wilderness Medical Society (WMS) Guidelines: The updated WMS Practice Guidelines for Spinal Cord Protection (2023–2024) provide strong, evidence-informed recommendations that directly bear on improvised collars. Key points include: rigid cervical collars (whether commercial or improvised) have “numerous identified risks and no demonstrated benefit” in wilderness trauma care.^8^ The guidelines do not require any cervical collar for most out-of-hospital scenarios; instead, they emphasize passive spinal motion restriction – i.e. encouraging a conscious patient to minimize neck movement and using soft padding or vacuum splints for support. They explicitly state that soft collars, commercial or improvised, may be used as one of several tools to reduce cervical motion as long as it does not interfere with critical care. Notably, a vacuum mattress or splint (if available) is preferred over any collar for spine protection, given its superior immobilization of the entire body and improved comfort. The WMS also notes there are no documented cases in the literature of neurologic harm from not using a collar, whereas there are plenty of reports of complications from collars. Thus, WMS suggests collars (even improvised) are optional adjuncts – helpful to remind patients not to move and to provide modest support, but not mandatory if careful handling is possible.

Military and EMS Protocols: Military field trauma care (TCCC guidelines) generally prioritize life-threatening hemorrhage control and airway management over spine immobilization, especially in active combat. Penetrating trauma with no neurologic signs is typically not immobilized at all. For blunt trauma in austere combat settings, if spinal injury is suspected, improvised immobilization may be used when tactical and logistical situations allow for example, a SAM splint or jacket may be applied during evacuation under fire if feasible. Formal studies or guidelines on specific improvised techniques in military literature are sparse; however, military medics are trained to be adaptable. The FastCast foam collar research noted above is a product of military orthopedic research aiming to improve austere care. Its superior performance in cadavers suggests that military teams might benefit from novel immobilization gear that is lightweight and conforms to the patient, potentially replacing the need to carry bulky C-collars in the future.

In civilian EMS, a landmark joint position statement by NAEMSP, ACS-COT, and ACEP in 2018 introduced the term “spinal motion restriction” and moved away from mandatory backboard + collar for all trauma. It advises that when SMR is indicated, a cervical collar (rigid) plus securing the patient supine on the stretcher is the usual method, but it also allows that if a collar cannot be applied or causes problems, manual stabilization or other adjuncts can be used. ^6,9^ Some EMS services in the U.S. and internationally have indeed shifted to soft collars or towel rolls with tape for select cases (particularly elderly patients or those who cannot tolerate rigid collars). The essence of these protocols is aligning with what wilderness practice has long had to do: tailor the immobilization to the situation, sometimes using creative measures to achieve relative stability without unduly harming or discomforting the patient.

## DISCUSSION

Improvised cervical spine immobilization can be effective in the wilderness, but its role should be understood in the broader context of spinal motion restriction and patient-centered care. The evidence reviewed indicates that several makeshift devices can significantly reduce neck motion, a necessary goal when a cervical injury is suspected, though none truly “immobilize” the spine completely. We discuss here the key considerations of biomechanical efficacy, field practicality, required training, patient safety, and limitations of improvised cervical collars, in light of our findings.

Biomechanical Efficacy: Studies on healthy volunteers show that improvised soft collars (fleece jackets, towel rolls) restrict cervical range of motion nearly as well as standard rigid collars in most directions.^1,10^ The 2019 WEM study and the 2022 LMIC trial both demonstrated non-inferiority of a rolled garment or towel, respectively, for limiting gross neck movements with the exception that extreme flexion may not be controlled as effectively by a towel.^1^ The SAM splint, a semi-rigid foam padded metal provides a firm structure akin to a medical collar and was shown to essentially match a Philadelphia collar’s stability.^11^ Importantly, the more recent cadaveric evidence (Roebke 2023) suggests that advanced improvised methods like a spray-on foam splint could even surpass traditional methods in truly unstable injuries.^2^ In summary, from a purely biomechanical perspective, improvisation is a valid strategy: a well-placed rolled jacket or molded splint can reduce cervical motion by roughly 50–75%, comparable to a commercial device. No device (including rigid collars) absolutely prevents all motion even rigid collars allow residual movement and can lose alignment under force.^6^ Thus, any collar should be viewed as an adjunct to careful patient handling rather than a fail-safe. The marginal differences in motion restriction between improvised and standard collars (often within a few degrees) suggest that in the wilderness context, achieving perfect immobilization is less realistic than focusing on overall spinal precautions (gentle extrication, avoiding unnecessary moves, keeping the head in neutral alignment).

Field Practicality: Improvised collars clearly excel in availability and adaptability. A fleece jacket or towel is almost always at hand in backcountry scenarios. These soft materials can be rapidly fashioned into a collar by rolling and wrapping around the neck, a technique that requires minimal equipment (perhaps just some tape or a pin to secure the roll).^12^ The process is intuitive: as one wilderness physician quips, “an improvised cervical collar can be made by firmly rolling and taping a fleece jacket around the head and neck”.^12^ SAM splints are also common in wilderness medical kits and, while a bit stiffer to work with, can be pre-shaped to approximate the contour of a neck. Practically, a SAM splint collar likely requires at least 2–3 straps or wraps (e.g. bandannas, belts, or duct tape) to hold its shape once applied; this is a bit more involved than a soft roll but offers more support. One advantage noted in the Porter et al. study was comfort subjects overwhelmingly found the improvised fleece more comfortable than a stiff collar.^10^ In the field, comfort is not a luxury; it can reduce patient movement (due to pain or anxiety) and improve cooperation during a prolonged evacuation. Soft materials conform to individual anatomy (important for patients with beards, occipital protuberances, etc.) and avoid pressure points. Given that wilderness evacuations often take hours, a comfortable improvised support that the patient can tolerate is arguably superior to an inflexible device that might cause discomfort or airway issues.

Weight and bulk are also practical considerations. Carrying a plastic cervical collar on every trip is impractical for most climbers or hikers, whereas a SAM splint weighs only a few ounces and is multipurpose. Clothing and towels double as normal gear. The spray-foam concept, if developed, comes in a small canister, a significant logistic benefit noted by its researchers.^2^ Environmental conditions in the wilderness can influence practicality as well. Rain or freezing conditions could affect improvised materials (a soaked towel may lose stiffness, a frozen jacket might become rigid). In general, materials like foam, down, or synthetic fill maintain some support when wet and can be augmented by waterproof layers if needed. There is little direct research on how improvised collars hold up in extreme environments, but anecdotal reports from mountain rescue indicate they can be used successfully even in alpine conditions by using what’s available (for example, wrapping a dry mid-layer clothing item around the neck even if outer clothes are wet).

Training Requirements: While improvisation encourages creative problem-solving, proper technique is still crucial to ensure efficacy and avoid harm. The studies reviewed mostly involved medical personnel or researchers applying the improvised devices under controlled conditions. For lay rescuers or minimally trained wilderness leaders, there is a learning curve. For instance, molding a SAM splint into a C-collar shape is a teachable skill wilderness first responder courses often include this in their curriculum but without prior practice, a rescuer might not achieve an optimal fit. Key training points include: ensuring the improvised collar is neither too loose (ineffective) nor too tight (impeding breathing or circulation), maintaining neutral alignment of the neck during application, and securing the device so it doesn’t shift during movement. In one training study (Hostler et al. 2005), EMS personnel practiced applying extrication collars and demonstrated the need for periodic retraining to maintain competence.^13^ We extrapolate that improvised collars similarly require hands-on practice to be done swiftly and correctly.

The good news is that improvisation techniques are widely taught in wilderness medicine programs. As illustrated in the NOLS case, even a fairly improvised setup (jacket collar + stuff-sack head bolsters) was applied well enough that EMS professionals accepted it and just replaced the collar with a soft one for transport.^6^ This suggests that the average trained wilderness first responder can indeed apply improvised SMR effectively. Military medics likewise receive training in resource-limited trauma care, which includes makeshift splinting. We found no specific study quantifying application times or error rates for improvised vs commercial collars, which is an area for future research. It would be useful to know, for example, if a team can apply a towel collar faster or slower than a rigid collar, and what common mistakes occur (improper positioning, inadequate securing, etc.). Until such data exist, training programs should continue emphasizing scenario-based practice: e.g., have students attempt to immobilize a mock patient’s neck with only the contents of a backpack. Mastery of these improvisation skills can make a critical difference when faced with an actual wilderness spine injury.

Patient Safety and Comfort: Safety considerations revolve around avoiding the known downsides of cervical collars while still protecting the spine. Improvised collars, by virtue of being softer and less constrictive, inherently mitigate some risks associated with rigid collars. For example, a soft roll will not significantly raise intracranial pressure but a rigid collar can impede venous drainage from the head, whereas a loosely fitted towel or jacket generally does not cause jugular vein compression.^6^ Airway management is also safer: a hard collar limits jaw opening and can complicate intubation or cause aspiration if the patient vomits; an improvised soft collar can be rapidly removed or adjusted if needed, and often leaves more room for airway interventions.

Pressure ulcers on the neck (sometimes seen with prolonged rigid collar use) are unlikely with a padded garment. The WMS guidelines note that any collar can conceal life-threatening signs (like neck vein distension or tracheal deviation),^8^ but an improvised collar (since it can be quickly undone) poses less of a barrier to repeated examinations.

On the other hand, improvised devices must be applied carefully to avoid creating new hazards. Duct tape, if used, should never encircle the neck or adhere to skin there only across the forehead or used to secure padding to a backboard. There have been rare reports (in non-wilderness settings) of patients choking or obstructing airway due to poorly placed straps; thus, rescuers should ensure no improvised support is pressing on the throat. Another safety aspect is maintenance during evacuation: an improvised collar might loosen or slip as the patient is moved over rough terrain. Periodic re-checks and adjustments are necessary. This is where a device like the FastCast foam shows promise once hardened in place, it didn’t shift at all under stress in the cadaver tests.^2^ Traditional improvised wraps may need additional support (e.g., incorporating the patient’s torso or a backboard). Indeed, Eisner et al. recommended using the folded towel in combination with strapping the patient to a board, to compensate for any laxity in flexion support.^1^

It is worth emphasizing that patient comfort is not at odds with safety; in fact, it enhances it. A comfortable patient is less likely to move suddenly or become agitated. The fleece collar’s improved comfort in Porter’s study is a clear advantage.^10^ Patients in pain or shock might better tolerate a gentle padding than a stiff device. In long evacuations, providing some cushioning (even if it sacrifices a tiny degree of immobilization fidelity) likely prevents other complications such as anxiety, respiratory compromise, or non-compliance. The current trend, per WMS and others, is indeed to prefer “comfortable patient packaging” over rigid immobilization.^6^ Improvised collars align well with that philosophy.

Device Limitations: No improvised solution is perfect, and understanding limitations helps determine when and how to use them. Soft improvised collars (towels, clothing) cannot completely prevent motion particularly flexion (forward bending) and traction (lengthwise movement) of the cervical spine. If a patient is combative or unable to cooperate, a soft collar alone will not restrain the neck; additional manual stabilization or a full-body vacuum immobilizer would be needed. Moreover, improvised collars generally provide minimal axial support they do not significantly unload the weight of the head from the spine (something even rigid collars only partially do). Thus, if there is a grossly unstable fracture, any collar is just buying time and motion reduction, not truly “splinting” the bone ends. Only careful handling and perhaps spinal motion restriction of the whole body can prevent exacerbation in such cases.

The security of improvised devices is another limitation. A SAM splint collar, if not securely fastened, might uncoil at inopportune moments. Tape adhesive could fail if wet or cold. A jacket used as a collar might loosen as fabric compresses. In practice, rescuers often combine methods: e.g., use a jacket collar and tie the patient’s head to a sleeping pad or use a bandage to further stabilize the head against the torso. These hybrids can be very effective (and are essentially a soft variant of the old sandbag-and-tape method). However, combining devices means more complexity and something that can potentially go wrong if not monitored (a knot slipping, etc.).

Another limitation is durability over time. A patient with an improvised collar who must be evacuated over many hours (or days, in remote expeditions) may experience changes: the collar could become soiled, wet, or may cause heat discomfort. Conversely, in cold weather, bulky improvised wraps might make it harder to insulate or could contribute to heat loss if not done carefully (though generally wrapping neck with clothing would increase warmth). Rescue teams should be prepared to modify the immobilization as needed en route e.g., replacing a soaked towel with a dry garment, or tightening straps after a bumpy carry.

Finally, it’s important to note that improvised collars have not been studied in pediatric patients specifically. Children have proportionally larger heads and more flexible spines, and the standard of care often uses padding under the shoulders to maintain neutral alignment. Improvised collars (like a small towel roll) are commonly recommended for children if a rigid pediatric collar isn’t available.^4^ But one must ensure the size is appropriate, a roll that’s too big could hyperextend a small child’s neck, for example. Thus, improvisation in pediatrics should be approached with particular caution and gentleness.

Integration with Spinal Motion Restriction: All sources agree that a collar alone (improvised or not) is not a standalone solution it’s one component of SMR. Proper spinal care in the wilderness also involves aligning the spine (bringing the patient’s head into a neutral position if possible without pain), maintaining that alignment with body positioning (e.g., placing the patient supine on a firm surface or vacuum mattress), and minimizing unnecessary movement (including during lifts or transport).^8^ An improvised collar can help maintain alignment by filling space and reminding the patient to minimize neck motion, but equally critical is how the whole patient is handled. Techniques like the multi-person lift (“lift-and-slide” or BEAM lift) are preferred over log-rolling a patient, to reduce twisting of the spine.^8^ If available, securing the patient with straps (even improvised webbing or cords) to a makeshift litter or stretcher will greatly augment cervical precautions by limiting gross body shifts. In essence, the collar – improvised or rigid is just one piece of the puzzle. Wilderness providers have to prioritize global stability and not develop a false sense of security from a collar. This point is underscored by the fact that no collar prevents all movement and none has shown clear outcome benefit in isolation. ^6,8^

Patient Monitoring and Reassessment: A final aspect to discuss is that when using improvised immobilization, diligent monitoring is necessary. Since these devices aren’t standardized, rescuers should continuously reassess neurologic status (any changes in sensation, strength, etc.), check device placement, and ensure that the patient’s comfort is balanced with stability. If at any point the improvised collar seems to be causing distress (breathing difficulty, excessive pain), it should be modified or loosened – unlike a rigid collar, which sometimes crews are reluctant to remove even if it’s problematic. The flexibility in approach is actually a benefit of improvisation: because you invented the solution on the spot, you can also adapt it on the spot.

Limitations of Current Evidence: From a research standpoint, the evidence base is still relatively small. Only two studies specifically on improvised collars (fleece and towel) were identified in a 2025 scoping review of prehospital spinal care,^14^ versus dozens of studies on commercial collars. Most were on healthy volunteers; only Roebke’s was on actual unstable spines (cadaveric). We lack direct clinical trials and likely will for ethical reasons testing these devices in actual trauma patients. Thus, we infer efficacy primarily from motion studies, not from demonstrated prevention of neurologic injury. Fortunately, as noted, there are no known cases where absence of a rigid collar in wilderness led to worse outcomes,^8^ which provides some reassurance that the focus on motion restriction (whether via improvised means or not) is appropriate. Another gap is long-term comfort data: an improvised collar might be fine for an hour, but what about 8 hours? There’s room for field studies or case series documenting real wilderness evacuations using improvised SMR, including patient feedback and any complications.

## CONCLUSION

Improvised cervical spine immobilization techniques from soft collars fashioned out of clothing or towels to molded splints and novel foam devices play an important role in wilderness and austere trauma care. Evidence to date indicates that these improvisations can substantially limit cervical motion, often nearly as well as standard collars, while offering advantages in availability, adaptability, and patient comfort.^10,11^ In keeping with evolving practice guidelines, rigid immobilization is no longer the priority; instead, spinal motion restriction (SMR) with an emphasis on gentle handling and maintaining alignment is the goal.^6,8^ Improvised collars are valuable tools within this paradigm, they provide support and a reminder for caution without many of the downsides of hard collars.

However, successful use of makeshift collars requires knowledge and training to apply them correctly and safely. Rescuers should be familiar with techniques like rolling a jacket or shaping a SAM splint, and always be vigilant about airway and circulation when using any collar.

Whenever possible, improvised cervical support should be combined with overall patient immobilization (e.g., securing the body to a litter or using a vacuum mattress) to maximize stability. Importantly, if an improvised device cannot be applied or maintained (such as in high-angle technical rescues or when a patient must be moved urgently due to environment), emphasis should shift to manual stabilization and rapid evacuation rather than dogmatic application of a collar.

In summary, makeshift cervical collars whether a humble towel or a high-tech foam spray have proven capable of reducing neck motion in the wilderness. They enhance our toolkit for spine injuries when conventional equipment is lacking. Their proper use, aligned with current guidelines, can improve patient comfort and potentially safety during lengthy evacuations. As wilderness medical practice advances, we foresee greater acceptance of soft and improvised immobilization and further innovation (like the spray-on splint) tailored for remote medicine. Ongoing education and research will continue to refine these techniques, but current evidence supports that improvised spinal precautions, done thoughtfully, are both practical and biomechanically sound in the austere environment.^1,11^

## ETHICAL CONSIDERATION AND REPORTING

This study was a review of existing literature and did not involve any new research on human or animal subjects; therefore, institutional review board approval was not required. We adhered to the Preferred Reporting Items for Systematic Reviews and Meta-Analyses (PRISMA) 2020 guidelines throughout the design and reporting of this review. In accordance with PRISMA recommendations, a flow diagram of study selection is provided (Figure 1), and a completed PRISMA checklist was prepared to ensure transparent and complete reporting of our methods and findings.

## Data Availability

All data produced in the present work are contained in the manuscript

## ACKNOWLEDGMENTS

The author thanks Wilderness Medical Society peers for critical feedback.12

## FINANCIAL/MATERIAL SUPPORT

None.

## REFERENCES

1. Eisner ZJ, Delaney PG, Pine H, et al. Evaluating a novel, low-cost technique for cervical-spine immobilization for application in resource-limited LMICs: a non-inferiority trial. Spinal Cord. 2022;60(8):726–732. doi:10.1038/s41393-022-00764-3

2. Roebke AJ, Bates N, Jurenovich K, et al. Cervical spinal immobilization: a head-to-head comparison of a one-step spray-on foam splint versus structural aluminum malleable splint immobilization. Mil Med. 2023;188(9-10):e2987–e2991. doi:10.1093/milmed/usad081

3. Society for Academic Emergency Medicine. Spinal immobilization. Council of Emergency Medicine Residency Directors (CORD) CDEM Curriculum. Updated 2024. Accessed July 11, 2025. https://www.saem.org/about-saem/academies-interest-groups-affiliates2/cdem/for-students/online-education/m3-curriculum/group-traumatic-and-orthopedic-injuries/spinal-immobilization

4. University of Washington Harborview Medical Center. Cervical collar for c-spine stabilization. Pediatric Trauma — Initial Stabilization and Transfer: Module 1. Updated 2023. Accessed July 11, 2025. https://pedtrauma.uw.edu/edition-3/module-1-initial-stabilization-and-transfer-pediatric-trauma/slide-15.

5. Sundstrøm T, Asbjørnsen H, Habiba S, Sunde GA, Wester K. Prehospital use of cervical collars in trauma patients: a critical review. J Neurotrauma. 2014;31(6):531–540. doi:10.1089/neu.2013.3094.

6. National Outdoor Leadership School (NOLS). Case study 34: cervical spine injury management in the wilderness. Updated 2023. Accessed July 11, 2025. https://www.nols.edu/case-studies/case-study-34/?answer=view#casestudy_answer.

7. National Outdoor Leadership School (NOLS). Case study 3: improvised cervical spine stabilization. Updated 2023. Accessed July 11, 2025. https://www.nols.edu/case-studies/case-study-3/?answer=view#casestudy_answer.

8. Wilderness Medical Society. 2024 spine management summary: evolving perspectives on spinal motion restriction. Wilderness Medicine Magazine. Published 2024. Accessed July 11, 2025. https://wms.org/magazine/magazine/1476/2024-Spine-Summary/default.aspx.

9. New Jersey Department of Health, Office of Emergency Medical Services. Spinal motion restriction: a position statement on prehospital care. Published 2018. Accessed July 11, 2025. https://www.nj.gov/health/ems/documents/education/smr_position_statement.pdf

10. Porter A, Difrancesca M, Slack S, Hudecek L, McIntosh SE. Improvised vs standard cervical collar to restrict spine movement in the backcountry environment. Wilderness Environ Med. 2019;30(4):412–416. doi:10.1016/j.wem.2019.07.002.

11. Real First Aid. SAM splint cervical collar: effectiveness in limiting cervical spine movement. Updated 2023. Accessed July 11, 2025. https://www.realfirstaid.co.uk/samsplintcollar.

12. Furman E. Improvised medicine: lessons from the wilderness. EM Resident. Published 2021. Accessed July 11, 2025. https://www.emra.org/emresident/article/improvised-medicine-lessons-from-the-wilderness.

13. Hostler D, McEntire SJ, Reilly PM, Rogers FB. Acquiring and maintaining competence in the application of extrication cervical collars by a group of first responders. Published 2008. Accessed July 11, 2025. https://www.researchgate.net/publication/26322228

14. Laermans J, Singletary EM, Macneil F, et al. Spinal motion restriction for possible traumatic cervical spine injury: a scoping review. Cureus. 2025;17(5):e84393. doi:10.7759/cureus.84393

